# Prevalence and determinants of Skilled Birth Attendance Among Women Who Delivered in the Past 12 Months in Sene East District, Ghana: A Community-Based Cross-Sectional Study

**DOI:** 10.64898/2026.07.22.26358729

**Authors:** Maxwell Adjiabadek Akesem, Daniel Ankomah, Joseph Kubi Appiah, Elijah Wamale Nlamba, Cyprian Issahaku Dorgbetor

**Affiliations:** Ghana Health Service, Sene East District Health Directorate, Health Promotion Unit, Kajaji, Ghana; Ghana Health Service, Sene East District Health Directorate, Reproductive and Child Health Unit, Kajaji, Ghana; Ghana Health Service, Sene East District Health Directorate, Kojokrom Health Center, Kajaji, Ghana; Ghana Health Service, Builsa Municipal Health Directorate, Disease Control Unit, Sandema, Ghana; Chinese University of Hong Kong, Faculty of Medicine, JC School of Public Health and Primary Care, Hong Kong, China; Ghana Health Service, Nkoranza Municipal Health Directorate, Nutrition Unit, Nkoranza, Ghana

**Author notes:** Corresponding author Email addresses: MAA.

**Keywords:** Skilled birth attendance, maternal health, antenatal care, determinants, Ghana

## Abstract

**Background:** Skilled birth attendance is essential to reduce maternal and neonatal mortality. However, the use of skilled delivery services is still low in many rural areas of Ghana including the Sene East District. This study investigated the level and determinants of skilled birth attendance among women who delivered in the district in the last 12 months.

**Methods:** A community-based cross-sectional study was conducted among 430 women who delivered within the last 12 months. The respondents were selected from Child Welfare Clinics by simple random sampling. Data was collected using a structured questionnaire and analysed using logistic regression. Variables with p-values <0.05 at the bivariate level were included in the multivariable logistic regression model. Statistical significance was p < 0.05.

**Results:** The prevalence of skilled birth attendance among respondents was 64.9%. In a multivariable logistic regression model, women who had a previous delivery at home were less likely to deliver with skilled birth attendants during the most recent pregnancy than first-time mothers (AOR = 0.1, 95% CI: 0.02–0.41). Women who had delivered at a traditional birth attendant’s (TBA) place during their previous delivery had significantly lower odds of skilled birth attendance (AOR = 0.1, 95% CI: 0.01–0.44). On the other hand, women who had previously delivered in a health facility were six times more likely to use skilled birth attendants than first-time mothers (AOR = 6.3, 95% CI: 1.32–30.7). Also, mothers living more than 10 km from the nearest health facility were 70% less likely to use skilled birth attendance than mothers living within 5 km of a health facility (AOR = 0.3, 95% CI: 0.14–0.80).

**Conclusion:** The uptake of skilled birth attendance in Sene East District is suboptimal. Enhancing access to health facilities, strengthening antenatal care services and addressing barriers to skilled delivery may improve maternal health service utilization in the district.

## Background

Maternal health remains a serious public health concern on a global scale. Skilled birth attendants is defined by the World Health Organization (WHO) as qualified medical professionals, such as midwives, physicians, or nurses, who have undergone the necessary education and training to successfully manage typical (uncomplicated) pregnancies, childbirth, and the early postnatal period (World Health Organization 2023). Additionally, skilled birth attendants are prepared to recognize, treat, and appropriately refer any complications that may develop in both mothers and infants (World Health Organization 2023).

Global maternal health initiatives continue to place a high premium on ensuring that all women have access to skilled care during childbirth. According to estimates, 700,000 more medical professionals and roughly 24,000 more birthing facilities will be needed to guarantee that, by 2030, all women have access to professional care during childbirth (WHO et al. 2018).

Access to skilled care during childbirth is a critical intervention for reducing maternal and neonatal mortality. Complications associated with pregnancy and childbirth are often unpredictable and can occur rapidly during labour, delivery, or the postpartum period, requiring timely and appropriate medical attention. Evidence indicates that most maternal deaths are preventable when skilled care is available, particularly for major obstetric complications such as unsafe abortion, infection, obstructed labour, haemorrhage, and hypertension (World Health Organization 2023).

Global commitments to improving maternal health have been emphasized through major development frameworks. The United Nations introduced the Millennium Development Goals (MDGs) in 2000, which were succeeded by the Sustainable Development Goals (SDGs) in 2015. Target 3.1 of SDG 3, which focuses on guaranteeing healthy lives and promoting well-being for everyone, aims to lower the global maternal death ratio to less than 70 per 100,000 live births by 2030 (United Nations Department of Economic and Social Affairs 2023).

Most maternal deaths are caused by a small number of direct obstetric problems. The majority of maternal deaths worldwide are caused by five direct causes: haemorrhage, sepsis, unsafe abortion, obstructed labour, and hypertensive disorders (von Dadelszen and Magee 2017). If pregnant women had access to the required obstetric care, most of these deaths could be prevented.

Significant regional differences in access to skilled delivery care persist despite improvements in maternal health indicators worldwide. Overall trends in childbirth assistance indicate a slow but steady increase in SBAs’ participation. Millions of women still give birth without getting the care they need, especially those who live in rural areas and are from economically disadvantaged backgrounds.

Maternal and neonatal mortality are still major problems in Sub-Saharan Africa. The Sub-Saharan African region’s higher rates of maternal and neonatal mortality are mostly caused by inadequate healthcare facilities, accessibility and unwillingness by pregnant women to use available healthcare facilities for delivery due to cultural norms, religious convictions, or financial constraints (Samuel, Zewotir, and North 2021).

Ghana lowered financial barriers to maternal health services by implementing governmental actions to support pregnant women. Ghana increased the usage of maternal health services by implementing the Free Maternal Healthcare Policy within the National Health Insurance Scheme (Twum et al. 2018). This policy permits pregnant women to receive prenatal, postnatal, and delivery services without having to pay for them directly. Despite these efforts, Ghana has had difficulty meeting the internationally accepted standard of 70 maternal deaths per 100,000 live births over the period (Twum et al. 2018). The use of trained birth attendants is influenced by a number of factors, according to earlier studies. Several factors, including a shortage of SBAs, the high cost of delivery services, and inadequate care during childbirth, have impeded progress in improving delivery care (Al-Kamali 2023; Kibria et al. 2017).

Numerous studies have examined factors influencing the use of expert delivery services in Ghana. These studies have reported that factors such as transportation and distance to health facilities and staff attitudes towards service users, as well as women’s age and education, place of residence (rural/urban), parity, socioeconomic status, and antenatal care attendance and socioeconomic status of women are associated with the uptake of SBAs in Ghana (Adongo et al. 2024; Gudu and Addo 2017; Manyeh et al. 2017; Saaka and Akuamoah-Boateng 2020; Yangnuu, Ampon-Wireko, and Nkrumah 2025).

Studies on factors influencing the use of skilled birth attendants have been done in Ghana, but there is limited evidence from rural and hard-to-reach districts such as Sene East. Socio-demographic characteristics, cultural beliefs, health service accessibility and previous maternal healthcare experiences may differ by context and may influence women’s choice of place of delivery and use of skilled birth attendants. Sene East is a predominantly rural district with dispersed settlements and limited access to some health services, thus posing unique maternal health challenges that may not be similar to those reported in urban or relatively accessible settings.

Moreover, available studies in Ghana have largely been based on regional or urban populations, with little community-based evidence from rural districts such as Sene East. Therefore, understanding the specific factors associated with skilled birth attendance in the district is important for generating context-specific evidence to inform maternal health interventions, improve utilization of skilled delivery services, and support efforts to reduce maternal and neonatal morbidity and mortality. Thus, the purpose of this study is to determine the prevalence and factors that influence skilled birth attendance in Sene East District. The results will provide evidence to support targeted programs aimed at enhancing maternal health outcomes and increasing the use of skilled delivery services in Ghana’s rural areas.

## Methods

### Study design and setting

This study employed a community-based cross-sectional design for this research. The study was carried out in the Sene East District of Ghana. Communities in the district are dispersed throughout the mainland and island settlements along the Volta Lake and the Sene River. The district is primarily rural. The district’s health centres, Community-Based Health Planning and Services (CHPS) compounds, and referral centres provide maternal health services, including prenatal, birth, and postnatal care.

Women of reproductive age who had given birth within the 12 months prior to the survey were the study’s target population. This time frame was chosen to ensure a sufficient sample of recent deliveries while reducing recall bias.

### Study population

The study population consisted of women aged 15–49 years who had delivered within the last 12 months and were currently residing in the district at the time of data collection.

### Inclusion criteria

Women had given birth in the 12 months before commencement of the study. Women participated in the study after giving their informed consent.

### Exclusion criteria

Women who were unable to answer the questionnaire or who were seriously ill throughout the time of data collection.

### Sample size determination

The single population proportion formula for cross-sectional research was used to calculate the sample size:

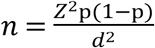

The computed sample size was as follows, assuming a 95% confidence level (Z = 1.96), a 5% margin of error (d = 0.05), and a 50% prevalence (p):

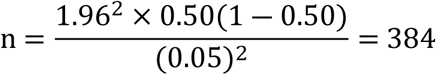

After adjusting for a 10% non-response rate, the final sample size was 422 participants. However, a total sample size of 430 participants was used in this study to increase the power of the study and to compensate for any unforeseen loss of data.

### Sampling procedure

A sample of women who had given birth in the previous 12 months was selected from Child Welfare Clinics (CWC) held at outreach and static points in the Sene East District. These clinics serve as points for children’s growth monitoring and immunization. Participants were chosen using the lottery method. Participants in the study were eligible women who visited the Child Welfare Clinics (CWC) during data collection. “Yes” and “No” were written on pieces of paper and put in a container and well mixed. Each qualified participant had to select one piece of paper from the container. Women who selected “Yes” were recruited and interviewed for the study, while those who selected “No” were not included. This process was repeated until the required sample size for the study was obtained.

### Study variables Dependent variable

Skilled birth attendance was the dependent variable. This variable was assessed using a binary result:

Yes– with the help of a trained birth attendant (nurse, physician assistant, doctor, or midwife). No– childbirth assisted by an untrained caregiver (self-delivery, a family member, or a traditional birth attendant).

### Independent variables

The independent variables were classified into three domains i.e. socio-demographic, obstetric and health system factors. Socio-demographic variables included maternal age, educational level, marital status, and occupation. Obstetric factors included parity (number of births), number of antenatal care (ANC) visits and place of delivery. Health system factors included distance to the health facility and the attitude of health care providers.

### Data collection tools and procedures

A standardized questionnaire was used to gather data from eligible women. The questionnaire was developed based on data from prior research on the use of maternal health services.

To ensure precise and effective data collection, Kobo Toolbox was used on mobile devices.

### Data analysis

Stata version 18 was used to analyse data that was exported from Kobo Toolbox.

The characteristics of the participants were summed up using descriptive statistics. Categorical variables were described using percentages and frequencies. Binary logistic regression was used in a bivariate analysis to evaluate the relationship between independent variables and skilled birth attendance. Variables with p-values <0.05 in the bivariate analysis were included in the multivariable logistic regression model to identify independent predictors of skilled birth attendance. Adjusted odds ratios (AORs) with 95% confidence intervals (CIs) were calculated, and statistical significance was set at p < 0.05.

### Ethical considerations

The Committee on Human Research, Publication, and Ethics (CHRPE) of Kwame Nkrumah University of Science and Technology (KNUST) granted ethical approval under approval number CHRPE/AP/609/26. Permission to conduct the study at several CWC centres was also given by the Sene East District Health Directorate. Each participant provided informed consent prior to data collection. The study was completely voluntary, and participant data was kept private.

## Results

### Socio-demographic characteristics of the respondents

The socio-demographic characteristics of the study participants are presented in Table 1. The study had 430 women who participated in it. The majority of respondents were between 25 to 29 years (30.23%) followed by those between 30 to 34 years (23.72%), while respondents aged below 20 years constituted 10.23% of the study population. The majority were married respondents (66.28%). Concerning the educational status, 37.91% of the respondents had no formal education, 24.88% had attained primary education, and only 3.26% had tertiary education. In terms of occupation, 38.14% were traders/fish mongers, 28.14% were farmers and 16.74% were unemployed/housewives. Most respondents were Christians (73.26%) while 13.72% were practicing traditional religion.

**Table 1.**
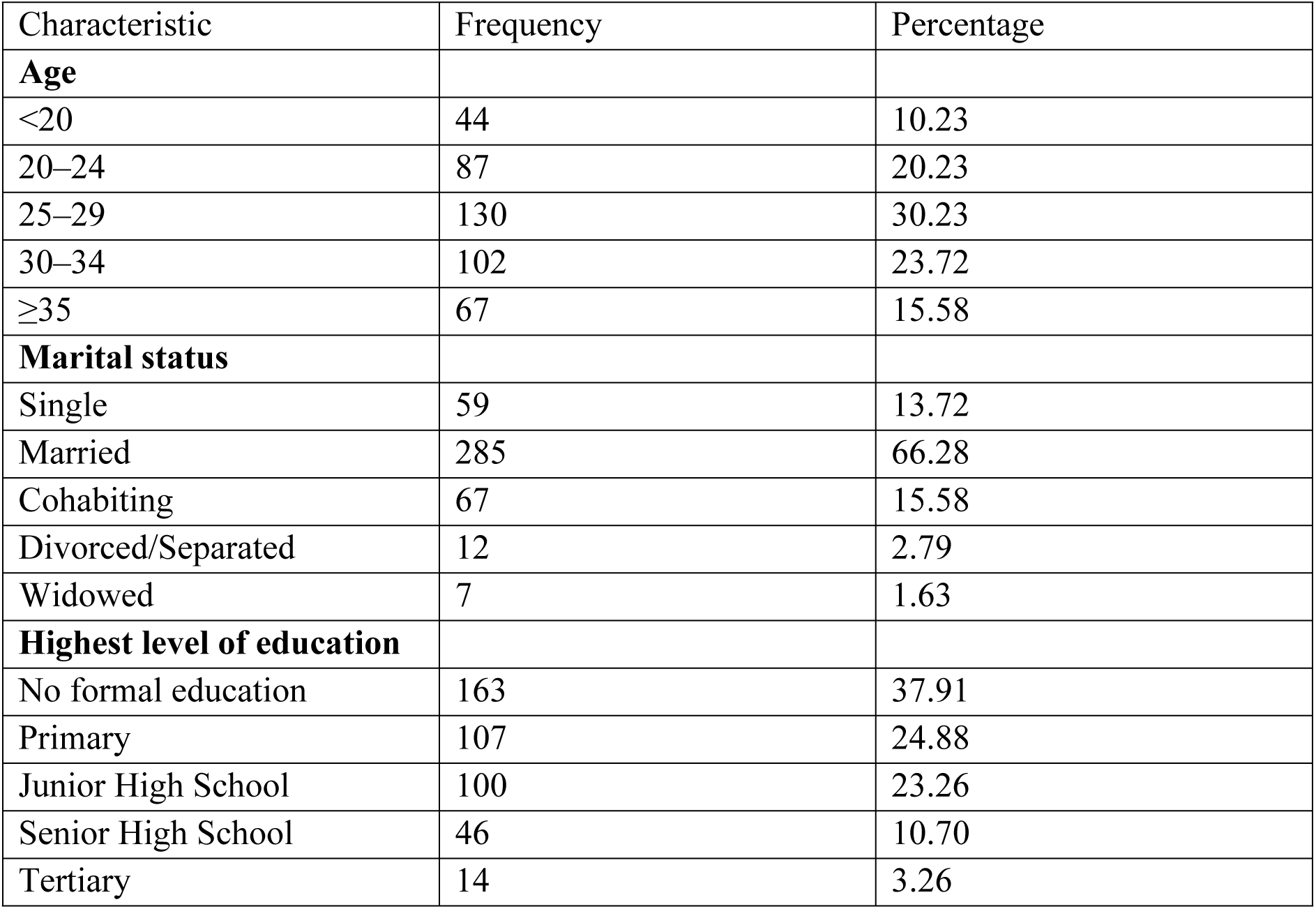
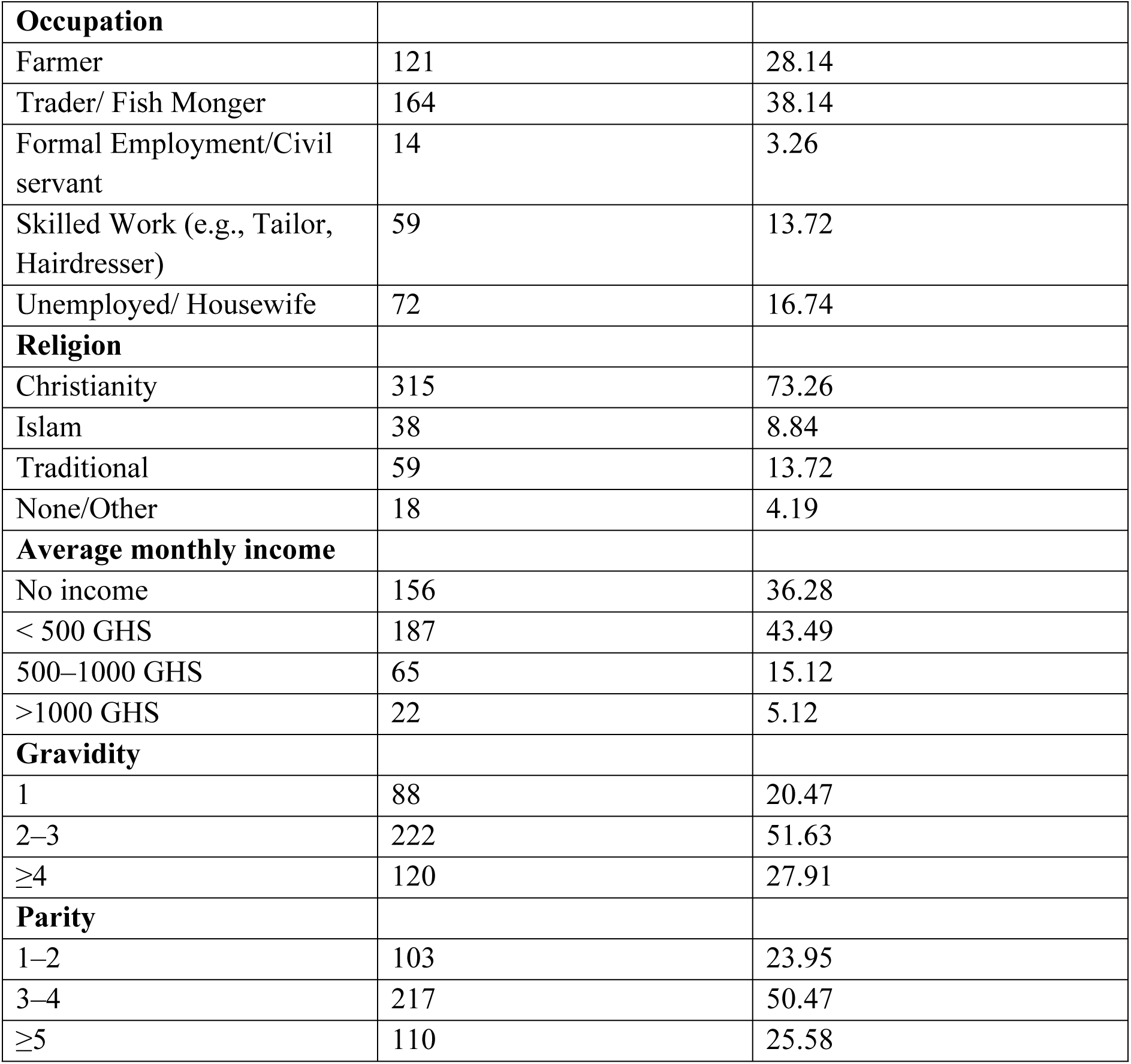
Socio-demographic characteristics of respondents.

### Bivariate analysis of factors associated with Skilled birth attendance

The bivariate logistic regression analysis showed several socio-demographic, obstetric, and health system factors that were associated with delivery by skilled birth attendants among women in Sene East District (Table 2).

**Table 2.** Factors associated with skilled birth attendance: bivariate logistic regression analysis.

| Variable | Delivery by SBAs |  | N | COR (95% CI) | P-value |
| --- | --- | --- | --- | --- | --- |
|  | No(N=151) | Yes(N=279) |  |  |  |
| **Age** |  |  |  |  |  |
| <20 | 8 | 36 | 44 | 1 |  |
| 20–24 | 28 | 59 | 87 | 0.4 (0.19–1.13) | 0.094 |
| 25–29 | 45 | 85 | 130 | 0.4 (0.17–0.97) | **0.045\*** |
| 30–34 | 45 | 57 | 102 | 0.2 (0.11–0.66) | **0.004\*** |
| ≥35 | 25 | 42 | 67 | 0.3 (0.14–0.92) | **0.034\*** |
| **Highest level of education** |  |  |  |  |  |
| No formal education | 76 | 87 | 163 | 1 |  |
| Primary | 39 | 68 | 107 | 1.5 (0.92–2.51) | 0.099 |
| JHS | 28 | 72 | 100 | 2.2 (1.31–3.83) | **0.003\*** |
| SHS/Tertiary | 8 | 52 | 60 | 5.6 (2.53–12.7) | **<0.001\*** |
| **Average monthly income** |  |  |  |  |  |
| No income | 50 | 106 | 156 | 1 |  |
| < 500 GHS | 73 | 114 | 187 | 0.7 (0.47–1.15) | 0.180 |
| 500–1000 GHS | 26 | 39 | 65 | 0.7 (0.38–1.28) | 0.258 |
| >1000 GHS | 2 | 20 | 22 | 4.7 (1.06–20.9) | **0.042\*** |
| **Who made the final decision on where to deliver?** |  |  |  |  |  |
| Woman alone | 35 | 31 | 66 | 1 |  |
| Partner alone | 18 | 31 | 49 | 1.9 (0.91–4.13) | 0.085 |
| Joint decision | 78 | 185 | 263 | 2.6 (1.54–4.64) | **<0.001\*** |
| Mother-in-law | 13 | 13 | 26 | 1.1 (0.45–2.79) | 0.793 |
| Other relative | 7 | 19 | 26 | 3.1 (1.13–8.26) | **0.027\*** |
| **Did your partner financially support your delivery?** |  |  |  |  |  |
| Yes | 122 | 245 | 367 | 1 |  |
| No | 29 | 34 | 63 | 0.5 (0.33–1.00) | 0.051 |
| **Did your partner accompany you to ANC at least once?** |  |  |  |  |  |
| Yes | 46 | 128 | 174 | 1 |  |
| No | 105 | 151 | 256 | 0.5 (0.33–0.78) | **0.002\*** |
| **Parity** |  |  |  |  |  |
| 1–2 | 28 | 75 | 103 | 1 |  |
| 3–4 | 70 | 147 | 217 | 0.7 (0.46–1.31) | 0.358 |
| ≥5 | 53 | 57 | 110 | 0.4 (0.22–0.71) | **0.002\*** |
| **Antenatal care (ANC) attendance during your last pregnancy?** |  |  |  |  |  |
| Yes | 139 | 270 | 409 | 1 |  |
| No | 12 | 9 | 21 | 0.38 (0.15–0.93) | **0.036\*** |
| **Previous delivery location (before last pregnancy)** |  |  |  |  |  |
| This was my first delivery (No previous birth) | 16 | 61 | 77 | 1 |  |
| Home | 108 | 25 | 133 | 0.1 (0.03–0.12) | **<0.001\*** |
| Health facility | 13 | 190 | 203 | 3.8 (1.74–8.41) | **<0.001\*** |
| TBA's place | 14 | 3 | 17 | 0.1 (0.01–0.21) | **<0.001\*** |
| **Distance to nearest health facility** |  |  |  |  |  |
| < 5 km | 38 | 111 | 149 | 1 |  |
| 5–10 km | 56 | 81 | 137 | 0.4 (0.29–0.81) | **0.006\*** |
| >10 km | 57 | 87 | 144 | 0.5 (0.31–0.85) | **0.011\*** |
| **How would you rate the health worker's attitude?** |  |  |  |  |  |
| Good | 98 | 214 | 312 | 1 |  |
| Fair | 45 | 63 | 108 | 0.6 (0.40–1.01) | 0.053 |
| Poor | 8 | 2 | 10 | 0.1 (0.02–0.54) | **0.007\*** |
| **Do you believe delivering in a health facility is safer?** |  |  |  |  |  |
| Yes | 136 | 277 | 413 | 1 |  |
| No | 15 | 2 | 17 | 0.1 (0.01–0.29) | **<0.001\*** |

SBA was significantly associated with maternal age. Women aged 25–29 years (COR = 0.4, 95% CI: 0.17–0.97), 30–34 years (COR = 0.2, 95% CI: 0.11–0.66) and 35 and above years (COR = 0.3, 95% CI: 0.14–0.92) had lower odds of delivering with skilled birth attendants compared to women aged below 20 years.

Maternal education was significantly and positively associated with skilled birth attendance. Women with JHS education were more likely to use skilled birth attendants than women with no formal education (COR = 2.2, 95% CI: 1.31–3.83). Similarly, women who had SHS/Tertiary education had significantly higher odds of skilled birth attendance (COR = 5.6, 95% CI: 2.53– 12.7). Monthly income was also associated with skilled birth attendance; women earning more than 1000 GHS monthly were more likely to deliver with skilled birth attendants compared with women with no income (COR = 4.7, 95% CI: 1.06–20.9).

Place of delivery decision-making was also significantly associated with skilled birth attendance. Women who made a joint decision with their partner on the place of delivery had higher odds of skilled birth attendance than their counterparts who decided alone (COR = 2.6, 95% CI: 1.54–4.64). Similarly, women who delivered with other family members were more likely to use a skilled birth attendant (COR=3.1, 95% CI: 1.13–8.26).

Partner involvement during pregnancy predicted skilled birth attendance. Women whose partners did not accompany them to antenatal care had lower odds of skilled birth attendance compared to those whose partners accompanied them at least once (COR = 0.5, 95% CI: 0.33– 0.78).

Skilled birth attendance was significantly associated with parity. Women with parity of five or more had decreased odds of giving birth with skilled birth attendants compared with women with parity of one to two (COR = 0.4, 95% CI: 0.22–0.71). Also, women who did not attend antenatal care during the last pregnancy had lower odds of skilled birth attendance compared to women who attended antenatal care (COR = 0.38, 95% CI: 0.15–0.93).

Previous place of delivery was highly associated with skilled birth attendance. Women who had previous home delivery (COR = 0.1, 95% CI: 0.03–0.12) and delivery at a traditional birth attendant’s place (COR = 0.1, 95% CI: 0.01–0.21) had significantly lower odds of skilled birth attendance as compared to first-time mothers. Conversely, women who had previously delivered in a health facility were more likely to have skilled birth attendance (COR = 3.8, 95% CI: 1.74–8.41).

Skilled birth attendance was significantly associated with distance to the nearest health facility. Women living within 5 km of a health facility were more likely to use skilled birth attendants than women living 5-10 km away (COR = 0.4, 95% CI: 0.29-0.81) and those living more than 10 km away (COR = 0.5, 95% CI: 0.31-0.85).

Women who rated the attitude of health workers as poor were less likely to use skilled birth attendants compared to those who rated the attitude of health workers as good (COR = 0.1, 95% CI: 0.02–0.54).

Perception about safety of facility delivery was significantly associated with skilled birth attendance. Women who did not believe that being delivered in a health facility is safer had significantly lower odds of using skilled birth attendants compared to those who believed that health facility delivery is safer (COR = 0.1, 95% CI: 0.01–0.29).

### Multivariable analysis

Multivariable logistic regression analysis was performed to ascertain the independent factors associated with delivery by skilled birth attendants among women in Sene East District, controlling for potential confounders (Table 3).

**Table 3.** Factors associated with skilled birth attendance: multivariable logistic regression analysis.

| Variable | Delivery by SBAs |  | N | COR (95% CI) | P-value |
| --- | --- | --- | --- | --- | --- |
|  | No(N=151) | Yes(N=279) |  |  |  |
| **Age** |  |  |  |  |  |
| <20 | 8 | 36 | 44 | 1 |  |
| 20–24 | 28 | 59 | 87 | 0.9 (0.26–3.51) | 0.972 |
| 25–29 | 45 | 85 | 130 | 0.4 (0.12–1.99) | 0.324 |
| 30–34 | 45 | 57 | 102 | 0.5 (1.11–2.36) | 0.394 |
| ≥35 | 25 | 42 | 67 | 1.6 (0.31–9.25) | 0.549 |
| **Highest level of education** |  |  |  |  |  |
| No formal education | 76 | 87 | 163 | 1 |  |
| Primary | 39 | 68 | 107 | 1.4 (0.62–3.39) | 0.382 |
| JHS | 28 | 72 | 100 | 1.3 (0.55–3.48) | 0.483 |
| SHS/Tertiary | 8 | 52 | 60 | 2.5 (0.58–10.9) | 0.214 |
| **Average monthly income** |  |  |  |  |  |
| No income | 50 | 106 | 156 | 1 |  |
| < 500 GHS | 73 | 114 | 187 | 0.7 (0.36–1.57) | 0.463 |
| 500–1000 GHS | 26 | 39 | 65 | 0.6 (0.24–2.00) | 0.503 |
| >1000 GHS | 2 | 20 | 22 | 1.7 (0.13–22.6) | 0.663 |
| **Who made the final decision on where to deliver?** |  |  |  |  |  |
| Woman alone | 35 | 31 | 66 | 1 |  |
| Partner alone | 18 | 31 | 49 | 1.5 (0.43–5.37) | 0.508 |
| Joint decision | 78 | 185 | 263 | 1.3 (0.54–3.57) | 0.488 |
| Mother-in-law | 13 | 13 | 26 | 0.6 (0.15–2.89) | 0.593 |
| Other relative | 7 | 19 | 26 | 0.7 (0.16–3.72) | 0.749 |
| **Did your partner accompany you to ANC at least once?** |  |  |  |  |  |
| Yes | 46 | 128 | 174 | 1 |  |
| No | 105 | 151 | 256 | 0.6 (0.31–1.23) | 0.171 |
| **Parity** |  |  |  |  |  |
| 1–2 | 28 | 75 | 103 | 1 |  |
| 3–4 | 70 | 147 | 217 | 1.1 (0.26–5.00) | 0.857 |
| ≥5 | 53 | 57 | 110 | 0.8 (0.15–5.09) | 0.894 |
| **Antenatal care (ANC) attendance during your last pregnancy?** |  |  |  |  |  |
| Yes | 139 | 270 | 409 | 1 |  |
| No | 12 | 9 | 21 | 1.1 (0.28–4.05) | 0.907 |
| **Previous delivery location (before last pregnancy)** |  |  |  |  |  |
| This was my first delivery (No previous birth) | 16 | 61 | 77 | 1 |  |
| Home | 108 | 25 | 133 | 0.1 (0.02–0.41) | **0.002\*** |
| Health facility | 13 | 190 | 203 | 6.3 (1.32–30.7) | **0.021\*** |
| TBA's place | 14 | 3 | 17 | 0.1 (0.01–0.44) | **0.006\*** |
| **Distance to nearest health facility** |  |  |  |  |  |
| < 5 km | 38 | 111 | 149 | 1 |  |
| 5–10 km | 56 | 81 | 137 | 0.4 (0.19–1.00) | 0.052 |
| >10 km | 57 | 87 | 144 | 0.3 (0.14–0.80) | **0.014\*** |
| **How would you rate the health worker's attitude?** |  |  |  |  |  |
| Good | 98 | 214 | 312 | 1 |  |
| Fair | 45 | 63 | 108 | 0.5 (0.27–1.30) | 0.195 |
| Poor | 8 | 2 | 10 | 0.2 (0.01–3.06) | 0.251 |
| **Do you believe delivering in a health facility is safer?** |  |  |  |  |  |
| Yes | 136 | 277 | 413 | 1 |  |
| No | 15 | 2 | 17 | 0.2 (0.03–1.32) | 0.097 |

The previous delivery site remained a strong predictor of skilled birth attendance. Women who had a previous delivery at home were less likely to deliver with skilled birth attendants during the most recent pregnancy than first-time mothers (AOR = 0.1, 95% CI: 0.02–0.41). Similarly, women who had delivered at a traditional birth attendant’s (TBA) place during their previous delivery had significantly lower odds of skilled birth attendance (AOR = 0.1, 95% CI: 0.01– 0.44). On the other hand, women who had previously delivered in a health facility were six times more likely to use skilled birth attendants than first-time mothers (AOR = 6.3, 95% CI: 1.32–30.7).

Distance to the nearest health facility remained significantly associated with skilled birth attendance after adjusting for other variables. Mothers who lived more than 10 km away from the nearest health facility were 70% less likely to use skilled birth attendance compared to mothers who lived within 5 km of a health facility (AOR = 0.3, 95% CI: 0.14–0.80). Mothers who lived 5–10 km away from a health facility were also less likely to use skilled birth attendance compared to those who lived within 5 km; however, this was not statistically significant (AOR = 0.4, 95% CI: 0.19–1.00).

Other variables such as maternal age, maternal educational level, monthly income, decision-making regarding place of delivery, partner accompanying to antenatal care, parity, antenatal care attendance, perceived health worker attitude and perception regarding safety of health facility delivery were not significantly associated with skilled birth attendance after adjusting for confounding factors.

## Discussion

The study assessed the prevalence and factors associated with skilled birth attendance among women who gave birth in the last 12 months in Sene East District. The study found that 64.9% of the women were assisted during delivery by a skilled birth attendant and 35.1% delivered without skilled assistance. The findings further revealed that there were several socio-demographic, obstetric and health system-related factors associated with skilled birth attendance among women in the district. Specifically, factors such as antenatal care attendance, maternal education, obstetric care, and accessibility to health facilities were significantly associated with the utilization of skilled birth attendants.

The study pointed out a substantial prevalence of skilled birth attendance, even though a considerable number of women in the district still give birth without the help of trained health professionals, despite interventions for maternal health. This has important public health implications as skilled birth attendance is one of the key strategies to reduce maternal and neonatal morbidity and mortality. The findings, therefore, provide important district-level evidence that could assist the Ghana Health Service and other stakeholders in designing targeted interventions to improve utilization of skilled delivery services especially in hard-to-reach rural communities within the district and similar settings in the country. The main findings of the study are discussed in the next sections in relation to the existing literature and possible contextual explanations.

Previous place of delivery was strongly associated with skilled birth attendance in both bivariate and multivariable analyses. First-time mothers had higher odds of using skilled birth attendants during the most recent delivery compared to women who had previously delivered at home or at a traditional birth attendant’s (TBA) home. Also, women who had delivered in a health facility in the past were more likely to use skilled birth attendants. Having delivered before in a health facility was associated with the likelihood of having skilled birth attendance for the most recent delivery. This finding is consistent with studies in Ethiopia and Eritrea that showed that previous exposure to facility-based delivery increases subsequent utilization of skilled birth attendants (Kifle et al. 2018; G. Tesfaye et al. 2019). Previous good experiences with health services can increase women’s confidence in health facilities and inform future decisions about delivery.

Distance to health facilities was an important factor influencing skilled birth attendance in this study. Women who lived >10 km from a health facility were less likely to use skilled birth attendants than women who lived within 5 km. This is in line with literature in Kenya, where it was found that women who lived closer to health facilities were more likely to use skilled delivery services (Ettarh and Kimani 2016; Gitimu et al. 2015) and in a multi-country analysis in Ghana, Kenya, Uganda and Tanzania, that increased distance was consistently associated with reduced skilled birth attendance (Adjiwanou and LeGrand 2013). Likewise, evidence from sub-Saharan Africa shows that the perception of distance as a major barrier is associated with higher use of unskilled delivery services (Addo et al. 2023).

Skilled birth attendance was strongly associated with maternal age, and women aged 25 years and above were less likely to use skilled birth attendants than women below 20 years. This implies that there is higher utilization of skilled delivery services among younger women and this decreases with increasing age. This finding is consistent with evidence from Ethiopia and the rest of sub-Saharan Africa that maternal age is a major determinant of skilled delivery use (Budu et al. 2021; Tesfaw et al. 2018). Similar findings have been reported in Nigeria and other low- and middle-income settings, showing that older age at first marriage is associated with a higher likelihood of skilled birth attendance (Afape et al. 2024). Maternal age is a consistent predictor of delivery care utilization alongside education, wealth and access to health services (Mezmur et al. 2017).

Monthly income was significantly associated with skilled birth attendance. Women who earned more than 1000 GHS per month were more likely to use skilled birth attendants than women with no income. This means that the higher the economic status, the higher the probability of using skilled delivery service. This finding is consistent with studies from Ethiopia and other settings that report household wealth as a key factor for skilled delivery utilisation (Mezmur et al. 2017; B. Tesfaye, Mathewos, and Kebede 2017). Similarly, studies from low- and middle-income countries have shown a positive association between wealth and skilled birth attendance (Wong et al. 2017). In line with this,(Houweling et al. 2007) also found that women in the poorest wealth categories are significantly less likely to use skilled delivery services than those in the richest groups.

Attendance of antenatal care (ANC) was significantly associated with skilled birth attendance, with women who did not attend ANC during their last pregnancy being less likely to use skilled birth attendants than women who attended ANC. The results indicate that ANC use strongly affects the use of skilled delivery. This finding is consistent with evidence from Ghana that ANC attendance is associated with significantly higher utilization of skilled delivery services (Gudu and Addo 2017; Manyeh et al. 2017). Ethiopia and Nigeria have reported similar results, indicating that increased ANC visits are associated with higher likelihood of skilled birth attendance (Mengesha et al. 2013; Okigbo and Eke 2015).

Maternal education was significantly associated with SBA; women with JHS and SHS/Tertiary education were more likely to use SBA as compared to women with no formal education. Skilled birth attendance was higher with an increased level of education. This is in line with studies in Ghana indicating a significant impact of maternal educational attainment on the utilization of skilled delivery services (Gudu and Addo 2017; Manyeh et al. 2017). Similar findings have been reported in Nigeria where women with secondary and higher education had higher odds of using skilled birth attendants than women without education (Afape et al. 2024). In Ethiopia, maternal education was also found to be an important predictor of skilled delivery service utilization (Mezmur et al. 2017). Similarly, a study in Kenya showed that mothers with tertiary or university education were significantly more likely to use skilled birth attendants during delivery than those with no education (Gitimu et al. 2015).

Parity was significantly associated with skilled birth attendance; women with five or more births were less likely to deliver with skilled birth attendants than women with one to two births. The findings suggest that as parity increases the use of skilled delivery decreases. The finding is consistent with studies in Ghana, Ethiopia and other sub-Saharan African countries that found that women of higher parity are less likely to use skilled birth attendants than women of lower parity (Manyeh et al. 2017; Moyer and Mustafa 2013; Teshale et al. 2020; Wilunda et al. 2015). Similarly, (Dankwah et al. 2019) found that primigravida women were more likely to deliver in health facilities than women who had previously given birth. Multiparous women had lower use of skilled birth attendants, which may reflect the perception of women with previous experience of childbirth that they are experienced and, thus, do not need a facility delivery, especially if previous deliveries were uncomplicated.

Partner involvement during pregnancy was significantly associated with skilled birth attendance. Women whose partners did not accompany them to antenatal care had lower odds of utilizing skilled birth attendants compared to women who were accompanied at least once. This means that male partner involvement in ANC has positive impact on skilled delivery attendance. This finding is consistent with studies from Ethiopia and sub-Saharan Africa which found that male partner involvement during ANC is associated with better facility delivery and skilled birth attendance (Degefa et al. 2024; Gessesse et al. 2024; Lateef et al. 2024). Similarly, (Teklesilasie and Deressa 2018) also found a strong and statistically significant association between husbands’ ANC involvement and women’s use of skilled attendants at birth.

Place of delivery decision-making was significantly associated with skilled birth attendance. Women who made decisions jointly with their partners were more likely to use skilled birth attendants compared with women who made decisions alone. Likewise, women who had made delivery decisions with family members had higher odds of skilled birth attendance. This finding is in line with studies conducted in Uganda, Eritrea, Ghana and other low- and middle-income countries that showed that the involvement of spouses or family members in delivery decision-making increases the likelihood of facility delivery and skilled birth utilization (Ameyaw et al. 2016; Anyait et al. 2012; Dankwah et al. 2019; Ganle et al. 2015; Kifle et al. 2018; Wilunda et al. 2015). In many contexts, the decision on where to deliver is shared or dominated by partners and family members and such involvement has been shown to facilitate access to skilled maternal care.

The perceived attitude of health workers was significantly associated with skilled birth attendance. Women who rated the attitude of health workers as poor were less likely to use skilled birth attendants than those who rated as good. This suggests that the interaction between provider and client is important in affecting delivery choices. This finding is consistent with the evidence from Ethiopia where disrespectful treatment from health workers has been identified as a major contributing factor for home delivery (Shiferaw and Modiba 2020). In Nepal, staff attitudes were also identified as an important determinant of skilled birth attendance (Raj et al. 2010). A systematic review also found that negative attitudes and behaviours of maternal health care providers are a global barrier to the utilization of maternal health services (Mannava et al. 2015).

Perception of safety of facility delivery was significantly associated with skilled birth attendance. Women who did not consider delivery in a health facility safer were less likely to use skilled birth attendants than those who considered facility delivery safer. This emphasizes the role of perceived safety on maternal health service utilization. This is consistent with qualitative evidence from Sierra Leone where women generally acknowledged the increased safety of facility delivery (Theuring, Koroma, and Harms 2018). In Nigeria too, the main reason for delivering babies in health facilities was perceived safety, as health facilities were seen as safer because of the presence of skilled health workers, ability to manage complications and better hygiene conditions (Hill et al. 2020).

### Limitations

There are some limitations of the study that need to be considered while interpreting the results. First, the study is cross-sectional, making it impossible to determine the causal relationships between the independent variables and skilled birth attendance. Consequently, the detected associations cannot be considered as causal relationships. Secondly, the study was based on self-reported information of the respondents, which is likely to be subject to recall bias with respect to past experiences of pregnancy and delivery. However, this was mitigated by limiting the study to women who had delivered in the last 12 months. Also, the study was conducted among women attending Child Welfare Clinics and it is possible that women who did not attend the clinics were not captured, which may affect the generalizability of the findings. Despite these limitations, this study provides important community-level evidence on the prevalence and determinants of skilled birth attendance in the Sene East District.

### Conclusion

The study demonstrated a suboptimal prevalence of skilled birth attendance in Sene East District amongst women who delivered in the last 12 months. Skilled birth attendance was significantly related to several socio-demographic, obstetric and health system-related factors. Skilled delivery utilization was particularly influenced by maternal education, antenatal care attendance, health facilities accessibility and previous delivery experiences. Our results underscore the need for targeted interventions to enhance access to maternal health services, improve antenatal care education, and overcome barriers to facility-based delivery in rural settings. Improving community-based maternal health education and access to skilled delivery services can help increase the use of skilled birth attendants and reduce maternal and neonatal morbidity and mortality.

## Declarations

### Ethics approval and consent to participate

Ethical approval was obtained from the Committee on Human Research, Publication, and Ethics (CHRPE) of Kwame Nkrumah University of Science and Technology (KNUST) with approval number CHRPE/AP/609/26. The study was conducted in accordance with the ethical principles outlined in the Declaration of Helsinki. Information on the study was provided at the first section of the survey and potential respondents were required to give informed consent by signing the consent form given to them. Confidentiality was maintained as names were not required, and the data was accessible only to the researchers.

### Consent for publication

Not applicable

### Availability of data and materials

The datasets used and analyzed during the current study are available from the corresponding author on reasonable request.

### Competing interests

The authors declare that they have no competing interests.

### Funding

This research received no specific grant from any funding agency

### Authors’ contributions

MAA, DA and JKA conceived the study, designed the methodology and coordinated data collection. MAA and EWN performed the statistical analysis and interpreted the results. MAA and DA drafted the manuscript. CID critically reviewed and revised the manuscript. All authors read and approved the final manuscript.

## Acknowledgements

The authors would like to express sincere gratitude to all respondents who participated in this study. Appreciation is also extended to the Sene East District Health Management Team and health facility staff, especially Assah Simon, Benjamin Osei Gyimah, Kumi Emmanuel, David

Fiagbor, David Nsoh, Sam Emmanuel, Kenneth Akayati, Jacob Yombo and Owusu Paul for facilitating the data collection process.

## Notes

### Competing Interest Statement

The authors have declared no competing interest.

## Reference

Addo, Isaac Yeboah, Evelyn Acquah, Samuel H Nyarko, Ebenezer N K Boateng, and Kwamena Sekyi Dickson. 2023. “Factors Associated with Unskilled Birth Attendance among Women in Sub-Saharan Africa: A Multivariate-Geospatial Analysis of Demographic and Health Surveys.” PLOS ONE 18(2): e0280992.

Adjiwanou, Vissého, and Thomas LeGrand. 2013. “Does Antenatal Care Matter in the Use of Skilled Birth Attendance in Rural Africa: A Multi-Country Analysis.” Social Science & Medicine 86: 26–34. 10.1016/j.socscimed.2013.02.047.

Adongo, Awinaba Amoah, Jonathan Mensah Dapaah, Francess Dufie Azumah, and John Nachinaab Onzaberigu. 2024. “Maternal and Child Health Care Access to Skilled Delivery Services among Ghanaian Rural Mothers.” Research in Health Services & Regions 3(1): 6. doi:10.1007/s43999-024-00042-0.

Afape, Ayobami Oyekunle, Precious Chidozie Azubuike, Oluwafunmilayo Oluwadamilola Ibikunle, and Amadou Barrow. 2024. “Prevalence and Determinants of Skilled Birth Attendance among Young Women Aged 15–24 Years in Northern Nigeria: Evidence from Multiple Indicator Cluster Survey 2011 to 2021.” BMC Public Health 24(1): 2471. doi:10.1186/s12889-024-19976-8.

Al-Kamali, Salwa. 2023. “Factors Influencing Access to and Utilization of Maternal Health Services in South Sudan.”

Ameyaw, Edward Kwabena, Augustine Tanle, Kwaku Kissah-Korsah, and Joshua Amo-Adjei. 2016. “Women’s Health Decision-Making Autonomy and Skilled Birth Attendance in Ghana.” International journal of reproductive medicine 2016(1): 6569514.

Anyait, Agnes, David Mukanga, George Bwire Oundo, and Fred Nuwaha. 2012. “Predictors for Health Facility Delivery in Busia District of Uganda: A Cross Sectional Study.” BMC Pregnancy and Childbirth 12(1): 132. doi:10.1186/1471-2393-12-132.

Budu, Eugene, Vijay Kumar Chattu, Bright Opoku Ahinkorah, Abdul-Aziz Seidu, Aliu Mohammed, Justice Kanor Tetteh, Francis Arthur-Holmes, Collins Adu, and Sanni Yaya. 2021. “Early Age at First Childbirth and Skilled Birth Attendance during Delivery among Young Women in Sub-Saharan Africa.” BMC Pregnancy and Childbirth 21(1): 834. doi:10.1186/s12884-021-04280-9.

von Dadelszen, Peter, and Laura A Magee. 2017. “Strategies to Reduce the Global Burden of Direct Maternal Deaths.” Obstetric medicine 10(1): 5–9.

Dankwah, Emmanuel, Wu Zeng, Cindy Feng, Shelley Kirychuk, and Marwa Farag. 2019. “The Social Determinants of Health Facility Delivery in Ghana.” Reproductive Health 16(1): 101. doi:10.1186/s12978-019-0753-2.

Degefa, Nega, Aster Dure, Dinkalem Getahun, Zekarias Bukala, and Tariku Bekelcho. 2024. “Male Partners Involvement in Their Wives’ Antenatal Care and Its Associated Factors in Southern Ethiopia. A Community-Based Cross-Sectional Study.” Heliyon 10(7): e28276. 10.1016/j.heliyon.2024.e28276.

Esena, Reuben K, and Mary-Margaret Sappor. 2013. “Factors Associated with the Utilization of Skilled Delivery Services in the Ga East Municipality of Ghana Part 2: Barriers to Skilled Delivery.” Int J Sci Tech Res 2(8): 195–207.

Ettarh, Remare R, and James Kimani. 2016. “Influence of Distance to Health Facilities on the Use of Skilled Attendants at Birth in Kenya.” Health Care for Women International 37(2): 237–49. doi:10.1080/07399332.2014.908194.

Ganle, John Kuumuori, Bernard Obeng, Alexander Yao Segbefia, Vitalis Mwinyuri, Joseph Yaw Yeboah, and Leonard Baatiema. 2015. “How Intra-Familial Decision-Making Affects Women’s Access to, and Use of Maternal Healthcare Services in Ghana: A Qualitative Study.” BMC pregnancy and childbirth 15(1): 173.

Gessesse, Nigusu Ayalew, Getahun Belay Gela, Amlaku Mulat Aweke, Fentahun Yenealem Beyene, Eden Asmare Kassahun, Alemwork Abie Getu, Bezawit Abeje Alemayehu, et al. 2024. “Male Partners’ Involvement in Antenatal Care and Its Associated Factors in West-Central Ethiopia.” BMC Public Health 24(1): 3015. doi:10.1186/s12889-024-20502-z.

Gitimu, Anne, Christine Herr, Happiness Oruko, Evalin Karijo, Richard Gichuki, Peter Ofware, Alice Lakati, and Josephat Nyagero. 2015. “Determinants of Use of Skilled Birth Attendant at Delivery in Makueni, Kenya: A Cross Sectional Study.” BMC Pregnancy and Childbirth 15(1): 9. doi:10.1186/s12884-015-0442-2.

Gudu, William, and Bright Addo. 2017. “Factors Associated with Utilization of Skilled Service Delivery among Women in Rural Northern Ghana: A Cross Sectional Study.” BMC Pregnancy and Childbirth 17(1): 159. doi:10.1186/s12884-017-1344-2.

Hill, Zelee, Pauline Scheelbeek, Joanna Schellenberg, and Yashua Hamza. 2020. “‘Everything Is from God but It Is Always Better to Get to the Hospital on Time’: A Qualitative Study with Community Members to Identify Factors That Influence Facility Delivery in Gombe State, Nigeria.” Global Health Action 13(1): 1785735. doi:10.1080/16549716.2020.1785735.

Houweling, Tanja A J, Carine Ronsmans, Oona M R Campbell, and Anton E Kunst. 2007. “Huge Poor-Rich Inequalities in Maternity Care: An International Comparative Study of Maternity and Child Care in Developing Countries.” Bulletin of the World Health Organization 85(10): 745–54. doi:10.2471/blt.06.038588.

Kibria, Gulam Muhammed Al, Swagata Ghosh, Shakir Hossen, Rifath Ara Alam Barsha, Atia Sharmeen, and S M Iftekhar Uddin. 2017. “Factors Affecting Deliveries Attended by Skilled Birth Attendants in Bangladesh.” Maternal Health, Neonatology and Perinatology 3(1): 7.

Kifle, Meron Mehari, Hana Fesehaye Kesete, Hermon Tekeste Gaim, Goitu Seltene Angosom, and Michael Berhane Araya. 2018. “Health Facility or Home Delivery? Factors Influencing the Choice of Delivery Place among Mothers Living in Rural Communities of Eritrea.” Journal of Health, Population and Nutrition 37(1): 22. doi:10.1186/s41043-018-0153-1.

Lateef, Monsurat A, Desmond Kuupiel, Gugu G Mchunu, and Julian D Pillay. 2024. “Utilization of Antenatal Care and Skilled Birth Delivery Services in Sub-Saharan Africa: A Systematic Scoping Review.” International Journal of Environmental Research and Public Health 21(4): 440. doi:10.3390/ijerph21040440.

Mannava, Priya, Kelly Durrant, Jane Fisher, Matthew Chersich, and Stanley Luchters. 2015. “Attitudes and Behaviours of Maternal Health Care Providers in Interactions with Clients: A Systematic Review.” Globalization and health 11(1): 36.

Manyeh, Alfred Kwesi, David Etsey Akpakli, Vida Kukula, Rosemond Akepene Ekey, Solomon Narh-Bana, Alexander Adjei, and Margaret Gyapong. 2017. “Socio-Demographic Determinants of Skilled Birth Attendant at Delivery in Rural Southern Ghana.” BMC Research Notes 10(1): 268. doi:10.1186/s13104-017-2591-z.

Mengesha, Zelalem Birhanu, Gashaw Andargie Biks, Tadesse Awoke Ayele, Gizachew Assefa Tessema, and Digsu Negesse Koye. 2013. “Determinants of Skilled Attendance for Delivery in Northwest Ethiopia: A Community Based Nested Case Control Study.” BMC Public Health 13(1): 130. doi:10.1186/1471-2458-13-130.

Mezmur, Markos, Kannan Navaneetham, Gobopamang Letamo, and Hadgu Bariagaber. 2017. “Individual, Household and Contextual Factors Associated with Skilled Delivery Care in Ethiopia: Evidence from Ethiopian Demographic and Health Surveys.” PLOS ONE 12(9): e0184688.

Moyer, Cheryl A, and Aesha Mustafa. 2013. “Drivers and Deterrents of Facility Delivery in Sub-Saharan Africa: A Systematic Review.” Reproductive Health 10(1): 40. doi:10.1186/1742-4755-10-40.

Okigbo, Chinelo, and Ahizechukwu Eke. 2015. “Skilled Birth Attendance in Nigeria: A Function of Frequency and Content of Antenatal Care.” African journal of reproductive health 19: 25–33.

Raj, Yuba, K Lyons, JVTE Skinner, and E R Van Teijlingen. 2010. “Determinants of Skilled Birth Attendants for Delivery in Nepal.” Kathmandu University Medical Journal 8(31): 325–32.

Saaka, Mahama, and Jones Akuamoah-Boateng. 2020. “Prevalence and Determinants of Rural-Urban Utilization of Skilled Delivery Services in Northern Ghana.” Scientifica 2020. doi:10.1155/2020/9373476.

Samuel, Oduse, Temesgen Zewotir, and Delia North. 2021. “Decomposing the Urban–Rural Inequalities in the Utilisation of Maternal Health Care Services: Evidence from 27 Selected Countries in Sub-Saharan Africa.” Reproductive Health 18(1): 1–12. doi:10.1186/s12978-021-01268-8.

Shiferaw, Biruhtesfa Bekele, and Lebitsi Maud Modiba. 2020. “Why Do Women Not Use Skilled Birth Attendance Service? An Explorative Qualitative Study in North West Ethiopia.” BMC Pregnancy and Childbirth 20(1): 633. doi:10.1186/s12884-020-03312-0.

Teklesilasie, Wondwosen, and Wakgari Deressa. 2018. “Husbands’ Involvement in Antenatal Care and Its Association with Women’s Utilization of Skilled Birth Attendants in Sidama Zone, Ethiopia: A Prospective Cohort Study.” BMC Pregnancy and Childbirth 18(1): 315. doi:10.1186/s12884-018-1954-3.

Tesfaw, Nigus, Ayu Gizachew, Getachew Mullu Kassa, and Amanuel Alemu Abajobir. 2018. “Skilled Delivery Service Utilization and Associated Factors among Mothers Who Gave Birth in the Last Two Years in Northwest Ethiopia.” Ethiopian journal of health sciences 28(4): 423–32. doi:10.4314/ejhs.v28i4.8.

Tesfaye, Brook, Tsedeke Mathewos, and Mihiretu Kebede. 2017. “Skilled Delivery Inequality in Ethiopia: To What Extent Are the Poorest and Uneducated Mothers Benefiting?” International Journal for Equity in Health 16(1): 82. doi:10.1186/s12939-017-0579-x.

Tesfaye, Gezahegn, Catherine Chojenta, Roger Smith, and Deborah Loxton. 2019. “Predisposing, Enabling and Need Factors Associated with Skilled Delivery Care Utilization among Reproductive-Aged Women in Kersa District, Eastern Ethiopia.” Reproductive Health 16(1): 167. doi:10.1186/s12978-019-0829-z.

Teshale, Achamyeleh Birhanu, Adugnaw Zeleke Alem, Yigizie Yeshaw, Sewnet Adem Kebede, Alemneh Mekuriaw Liyew, Getayeneh Antehunegn Tesema, and Chilot Desta Agegnehu. 2020. “Exploring Spatial Variations and Factors Associated with Skilled Birth Attendant Delivery in Ethiopia: Geographically Weighted Regression and Multilevel Analysis.” BMC Public Health 20(1): 1444. doi:10.1186/s12889-020-09550-3.

Theuring, Stefanie, Alimamy Philip Koroma, and Gundel Harms. 2018. “‘In the Hospital, There Will Be Nobody to Pamper Me’: A Qualitative Assessment on Barriers to Facility-Based Delivery in Post-Ebola Sierra Leone.” Reproductive Health 15(1): 155. doi:10.1186/s12978-018-0601-9.

Twum, Peter, Jing Qi, Kasangye Kangoy Aurelie, and Lingzhong Xu. 2018. “Effectiveness of a Free Maternal Healthcare Programme under the National Health Insurance Scheme on Skilled Care: Evidence from a Cross-Sectional Study in Two Districts in Ghana.” BMJ open 8(11): e022614. doi:10.1136/bmjopen-2018-022614.

United Nations Department of Economic and Social Affairs. 2023. “The Sustainable Development Goals Extended Report 2023 3 Good Health and Well-Being.” United Nations (April).

WHO, UNFPA, UNICEF, ICM, ICN, FIGO, and IPA. 2018. “Definition of Skilled Health Personnel Providing Care during Childbirth: The 2018 Joint Statement.” Who: 1–4. www.who.int.

Wilunda, Calistus, Gianluca Quaglio, Giovanni Putoto, Risa Takahashi, Federico Calia, Desalegn Abebe, Fabio Manenti, et al. 2015. “Determinants of Utilisation of Antenatal Care and Skilled Birth Attendant at Delivery in South West Shoa Zone, Ethiopia: A Cross Sectional Study.” Reproductive Health 12(1): 74. doi:10.1186/s12978-015-0067-y.

Wong, Kerry L M, María Clara Restrepo-Méndez, Aluísio J D Barros, and Cesar G Victora. 2017. “Socioeconomic Inequalities in Skilled Birth Attendance and Child Stunting in Selected Low and Middle Income Countries: Wealth Quintiles or Deciles?” PLOS ONE 12(5): e0174823.

World Health Organization. 2023. Trends in Maternal Mortality 2000 to 2020: Estimates by WHO, UNICEF, UNFPA, World Bank Group and UNDESA/Population Division. World Health Organization.

Yangnuu, Solomon, Sabina Ampon-Wireko, and Joseph Kwasi Nkrumah. 2025. “Factors Influencing the Utilization of Skilled Delivery Services in the Juaboso District of Western North Region, Ghana.” Journal of Global Health Science 7(2): 1–13. doi:10.35500/jghs.2025.7.e22.

